# Dengue NS1 interaction with lipids alters its pathogenic effects on monocyte derived macrophages

**DOI:** 10.1101/2024.05.24.24307786

**Authors:** Shashika Dayarathna, Bhagya Senadheera, Chandima Jeewandara, Madushika Dissanayaka, Farha Bary, Graham S. Ogg, Gathsaurie Neelika Malavige

## Abstract

**Background:** While dengue NS1 antigen has been shown to be associated with disease pathogenesis in some studies, it has not been linked in other studies, with the reasons remaining unclear. NS1 antigen levels in acute dengue are often associated with increased disease severity, but there have been a wide variation in results based on past dengue infection and infecting dengue virus (DENV) serotype. As NS1 engages with many host lipids, we hypothesize that the type of NS1-lipid interactions alters its pathogenicity.

**Methods:** Primary human monocyte derived macrophages (MDMs) were co-cultured with NS1 alone or with HDL, LDL, LPS and/or platelet activating factor (PAF) from individuals with a history of past dengue fever (DF=8) or dengue haemorrhagic fever (DHF=8). IL-1β levels were measured in culture supernatants, and gene expression analysis carried out in MDMs. Monocyte subpopulations were assessed by flow cytometry. Hierarchical cluster analysis with Euclidean distance calculations were used to differentiate clusters. Differentially expressed variables were extracted and a classifier model was developed to differentiate between past DF and DHF.

**Results:** Significantly higher levels of IL-1β were seen in culture supernatants when NS1 was co-cultured with LDL (p=0.01), but with lower levels with HDL (p=0.05). MDMs of those past DHF produced more IL-1β when NS1 with PAF (p=0.02). MDMs of individuals with past DHF, were significantly more likely to down-regulate *RPLP2* gene expression when macrophages were co-cultured with either PAF alone, or NS1 combined with PAF, or NS1 combined with LDL. When NS1 was co-cultured with PAF, HDL or LDL two clusters were detected based on *IL10* expression, but these did not differentiate those with past DF or DHF.

**Conclusions:** As RPLP2 is important in DENV replication and in regulating cellular stress responses and immune responses and IL-10 is associated with severe disease, it would be important to further explore how differential expression of RPLP2 and IL-10 could lead to disease pathogenesis based on NS1 and lipid interactions.

## Introduction

The dengue virus (DENV) is estimated to infect 390 million individuals annually, resulting in almost 100 million symptomatic/apparent dengue infections [6]. Although most individuals who are infected with the DENV develop inapparent or mild illness, a proportion develop severe disease manifestations due to plasma leakage, organ dysfunction and severe bleeding [25]. The reasons for occurrence of symptomatic and severe disease in some individuals remain incompletely understood. Many factors appear to be involved such as presence of host comorbidities (diabetes, cardiovascular disease and obesity), heterologous infections, the magnitude, breadth and the specificity of DENV specific antibody and T cell responses, the virulence of the infecting DENV strain, the extent viraemia and an altered dysfunctional immune responses [7,16,25,37].

The DENV infects many different cell types including monocytes, macrophages, hepatocytes and B cells causing disease pathogenesis [16,25]. However, in addition to the virus, dengue NS1 independently has shown to contribute to disease pathogenesis by disrupting the endothelial glycocalyx [4,32], inducing cytokine production from immune cells [26], activating phospholipase A2 enzymes [34] and immune evasion by interfering with complement activation [3], based on *in vitro* experiments and in dengue mouse models. However, studies in symptomatic mouse models using non-mouse adapted DENV-2 virus strains show that DENV-induced pathology was not ameliorated by passive transfer of NS1-specific antibodies or by prior vaccination with recombinant NS1 [22]. Furthermore, based on data from patients with acute dengue, the relative contribution of NS1 in causing disease pathogenesis and vascular leak has been questioned. For instance, circulating NS1 levels are higher in primary dengue, which is usually associated with a lower risk of severe dengue [13,38]. Furthermore, while high NS1 antigen levels have shown to associate with severe dengue, not all individuals with high levels develop severe disease [23]. Plasma leakage in patients with acute illness usually occurs between day 3 to 6 of illness when the NS1 levels decline [25].

It was recently shown that NS1 binds to HDL and with a lower affinity to LDL, and NS1-HDL complexes induced proinflammatory cytokine production from monocyte-derived macrophages *in vitro* [5]. While many early studies have suggested that secretory NS1 circulates as a hexamer, with a lipid core, more recent studies suggest that secretory NS1 circulates predominantly as a tetramer associated with HDL [11]. During the course of the illness, the apolipoproteins that associate with NS1 have shown to change, with ApoAI and ApoB interacting with NS1 during acute illness but replaced by ApoE towards the recovery phase of illness [5]. Therefore, the lipid composition of the NS1 hexamer/tetramer or lipids that interact with NS1, could also contribute to the virulence of NS1 and potentially help explain some of the variability in previous studies [40]. Given the timing of vascular leakage that occurs in acute dengue and due to the varied data on NS1 antigen levels and disease severity in acute dengue, it is possible that the NS1 interactions with lipid could alter its pathogenicity, during the course of illness.

We have shown that many inflammatory lipid mediators such as platelet activating factor (PAF), leukotrienes, phospholipase A2 enzymes and prostaglandins metabolites are elevated in patients with acute dengue [15,18,19,35]. In mouse models, DENV infection was shown to induce intestinal inflammation, leading to gut barrier dysfunction [30] also observed in patients with acute dengue [10,33,39]. HDL is known to bind to LPS, and dysfunctional HDL (HDL with altered surface proteins and lipid cargo) was associated severe disease outcomes in sepsis [27,36]. It is possible that LPS binds to NS1, to increase the pathogenic effect of NS1 and we previously showed that *in vitro* LPS and the DENV acted synergistically to increase proinflammatory cytokine production [20]. However, the monocyte responses to the DENV varied widely between individuals, with those with a past history of dengue haemorrhagic fever (DHF) producing more virus and inflammatory mediators in their culture supernatants [21]. Therefore, to investigate if the pathogenicity of NS1 is altered when it interacts with different lipid mediators, we studied their effects on primary monocyte derived macrophages in individuals with a past history of mild or asymptomatic dengue and DHF.

## Methodology

### Culture of monocytes with different lipid mediators

Primary human monocytes were isolated from peripheral blood mononucleocytes (PBMCs) from 16 healthy individuals with past inapparent dengue/DF (=8) or DHF (n=8). Those with a past history of DHF were who were hospitalized with an acute dengue infection and who were classified as having DHF based on the WHO 2011 guidelines [41]. Of those who had past DF/inapparent dengue, 3 were hospitalized due to a dengue infection, but had no evidence of DHF, one was treated as an outpatient and 4 had an inapparent dengue infection (Supplementary table 1). The four individuals who had an inapparent dengue infection, were found to have neutralizing antibodies to several DENV serotypes by the FRNT assay, suggestive of past multi-typic dengue infections.

Monocyte isolation was carried out using CD14 magnetic beads (Miltenyi Biotech, Germany) and cultured in 96 well cell culture plates (0.125 x10^6^ cells in 250µl media per well), for 7 days with 5% CO_2_ at 37°C to convert into macrophages. The resting media contained RPMI-1640 (Thermofisher Scientific, USA), 2 mM L-glutamine (Thermofisher Scientific, USA), 1% penicillin-streptomycin (Thermofisher Scientific, USA), 10 mM Na Pyruvate (Thermofisher Scientific, USA), 10 mM HEPES (Thermofisher Scientific, USA), 1% MEM vitamins (Thermofisher Scientific, USA), 1% NEAA (Thermofisher Scientific, USA), 50 lM beta-mercaptoethanol (Thermofisher Scientific, USA), and 15% AB-ve human serum (Sigma-Aldrich, USA). After 7 days, resting media was removed, and the monocyte derived macrophages co-cultured with DENV-1-NS1 antigen (NativeAntigen, USA) (2µg/ml), LPS (100ng/ml, Invivogen, USA), PAF (500ng/ml, Sigma Aldrich, USA), HDL (2.5µg/ml, Sigma Aldrich, USA) and LDL (20µg/ml, Sigma Aldrich, USA) alone or with NS1 in combination with the different lipid mediators. Macrophages co-cultured with media alone (5% FBS instead) was considered as the controls. The plates were rocked at every 15 minutes for 90 minutes for homogenous distribution of media and rested for 24 hours at 37°C with 5% CO2 after which the culture supernatants and cells were harvested and RNA extracted. IL1-β levels were measured in culture supernatants using a quantitative ELISA (R&D systems, USA, Cat no: DY201) according to manufacturer’s instructions.

### Ethics approval

Ethics approval was obtained from the Ethics Review Committee of the University of Sri Jayewardenepura. All participants gave informed written consent.

### RNA extraction and quantitative real time PCR

RNA was extracted using Geneaid rSYNC RNA Isolation kit (Geneaid Biotech Ltd, Taiwan, Cat No: RDHF100) and converted into cDNA using high-capacity cDNA Reverse Transcription kit with RNase inhibitor (Thermofisher Scientific, USA). Quantitative real-time PCR was carried out in triplicate for the relative expression of GAPDH, IL-10, NLRP3, IFN-β1, RIGI, RPLP2 and SMPD1 (Thermofisher Scientific, USA) using Invitrogen Superscript III Platinum master mix kit reagents (Thermofisher Scientific, USA) in ABI 7500 (Applied Biosystems, USA). ΔCt values were calculated for each gene for each test condition. ΔΔCt values for each gene was calculated separately by subtracting the endogenous GAPDH ΔCt values of each test condition followed by Fold change (FC) calculation for each gene for different test conditions Due to the limited availability of sample volume, Gene expression experiments were performed for 12/16 individuals (DF=6, DHF=6).

### Phenotyping of monocytes

To characterize the monocyte populations flowcytometry was carried out in whole blood of the 12 individuals in whom we performed gene expression analysis (DHF=6, DF=6). Briefly, red blood cell (RBC) lysis was carried out using RBC lysis buffer (BD Biosciences, USA) according to manufacturer’s protocol. Cells were stained with Live/Dead Fixable aqua (Invitrogen, USA), CD14-FITC (BD Biosciences, USA), CD16-Pacific blue (BD Biosciences, USA) and acquired through FACSAria III (BD Biosciences, USA). The results were analyzed in FlowJo version 10.8.1 (BD Biosciences, USA) and the gating strategy for different populations of monocytes is shown in supplementary figure 1.

### Statistical analysis

Statistical analysis was performed using GraphPad Prism version 10 and (Dotmatics, USA), and R software. Mann-Whitney U test was performed when comparing two unpaired samples, and the Wilcoxon test was used to compare paired samples. Hierarchical cluster analysis with Euclidean distance calculations were used to differentiate clusters under different culture conditions of NS1 with different lipid mediators. Clustering and visualization of heatmaps for gene expression under different culture conditions were performed with R V4.3.2. Differentially expressed variables were extracted based on two parameters (>0.1 FC and < 0.1 p-value) and a volcano plot was constructed. The conditions which satisfy both the above parameters were identified as potential features in differentiating DF from DHF and the results were visualized in a scatter plot. Based on the extracted features, a classifier model was developed to differentiate the biological groups with Random Forest decision tree (10-fold cross validation) [8]. Accuracy of the model was validated internally and externally (Confusion Matrix and Cohen Kappa).

## Results

### IL-1**β** levels in culture supernatants with NS1 and different lipid mediators

Significantly higher levels of IL-1β were seen in culture supernatants when NS1 was co-cultured with LDL (p=0.01), while lower IL-1β were detected when NS1 was co-cultured with HDL (p=0.05) and PAF (p=0.82) than with NS1 alone which appeared to have a suppressive effect (Figure 1A). The IL-1β levels in media alone, with NS1, LPS and NS1 and LPS were similar to the levels observed by Benfrid et al. [5]. Interestingly, when monocyte derived macrophages were co-cultured with PAF and NS1 they produced a significantly higher IL-1β level than when co-cultured with PAF alone (p<0.0001). Similarly, the macrophages produced significantly higher levels of IL-1β (p<0.0001) when NS1 was co-cultured with LDL than with LDL alone. In contrast, IL-1β levels were significantly lowered when NS1 was co-cultured with HDL compared to HDL (p=0.03) (Figure 1A).

**Figure 1.**
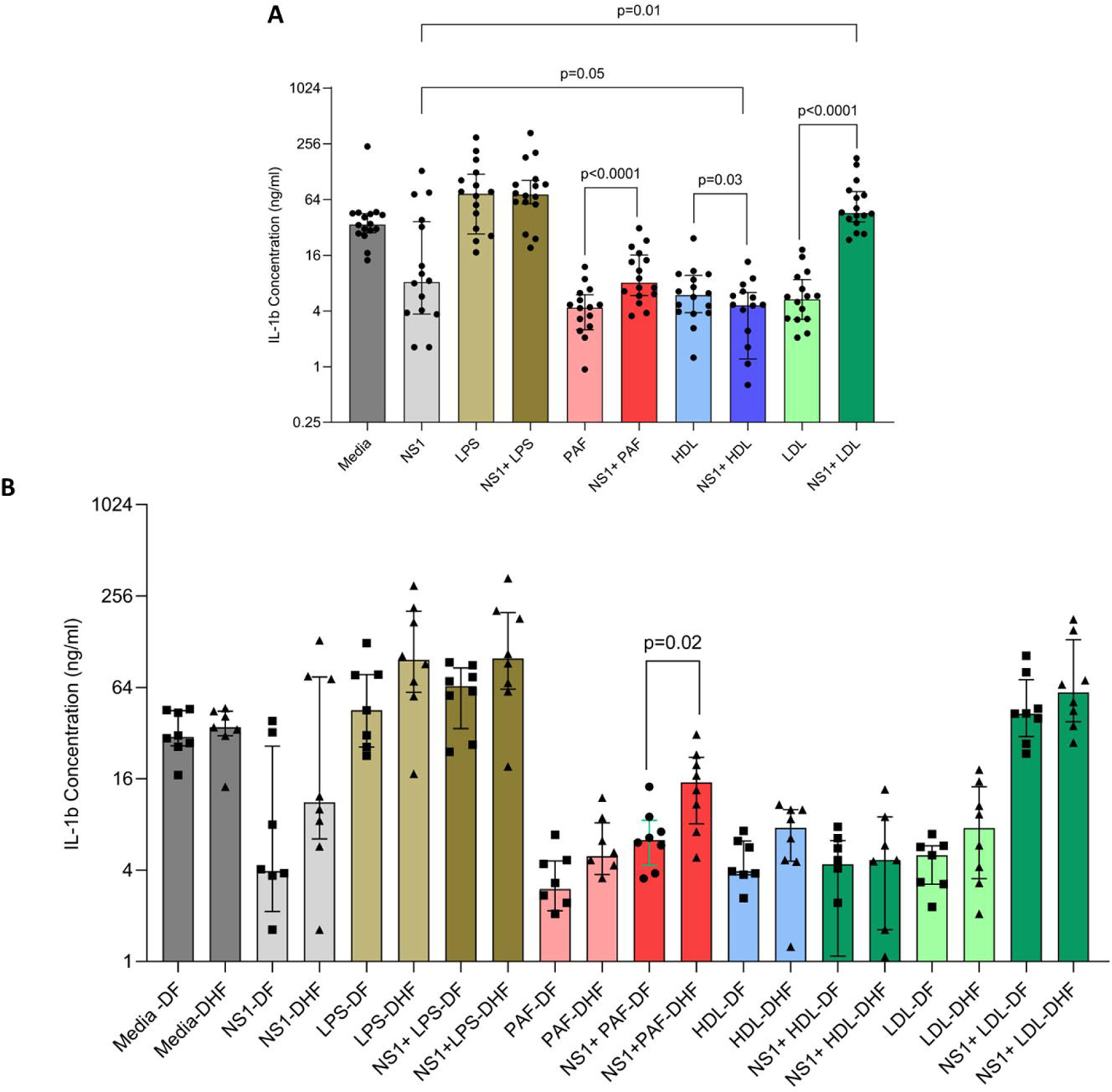
IL-1β levels in culture supernatant of primary human monocyte derived macrophages (MDMs) co-cultures with NS1 and different lipid mediators. MDMs from individuals with a history of DF (n=8) and DHF (n=8) were co-cultured with NS1 alone, different lipid mediators alone or with NS1 and different lipid mediators and the IL-1β levels in the supernatants were measured by ELISA after 24hrs. The Wilcoxon matched pairs signed rank test was used for comparison of IL-1 β levels in culture supernatants in the same individual under different conditions (A). IL-1β levels in culture supernatant in those with past DF vs DHF, under different culture conditions were compared using Mann-Whitney U test for unpaired groups (B). The lines indicate the median and IQR.

Previously we found that monocyte-derived macrophages of those with a past history DHF produced higher levels of cytokines and higher viral loads when infected with different DENV serotypes than in those with past DF [21]. Therefore, we sought to investigate if similar changes were seen when monocyte derived macrophages from individuals with past DF and DHF produced different levels of IL-1β under different conditions. We found that although those with past DHF produced a trend towards more IL-1β when NS1 was co-cultured with LPS and LDL, but differences were only significant when NS1 was co-cultured with PAF (p=0.02) (Figure 1B).

### Differential gene expression of monocyte-derived macrophages with NS1 co-cultured with different lipid mediators

We carried out hierarchical cluster analysis to identify differential expression of *IL-10, NLRP3, IFN-*β*1, RIGI, RPLP2* and *SMPD1* when NS1 was co-cultured with different lipid mediators. Accordingly, we only identified two main clusters based on the expression of *IL-10* (Figure 2). In cluster 1, high *IL-10* expression was seen when monocyte derived macrophages were co-cultured with PAF alone, NS1+PAF, NS1, LDL, HDL and NS1+HDL. Based on the overall expression of other genes, no significant differences were found. The two main clusters identified did not distinguish between those with past DF or DHF.

**Figure 2.**
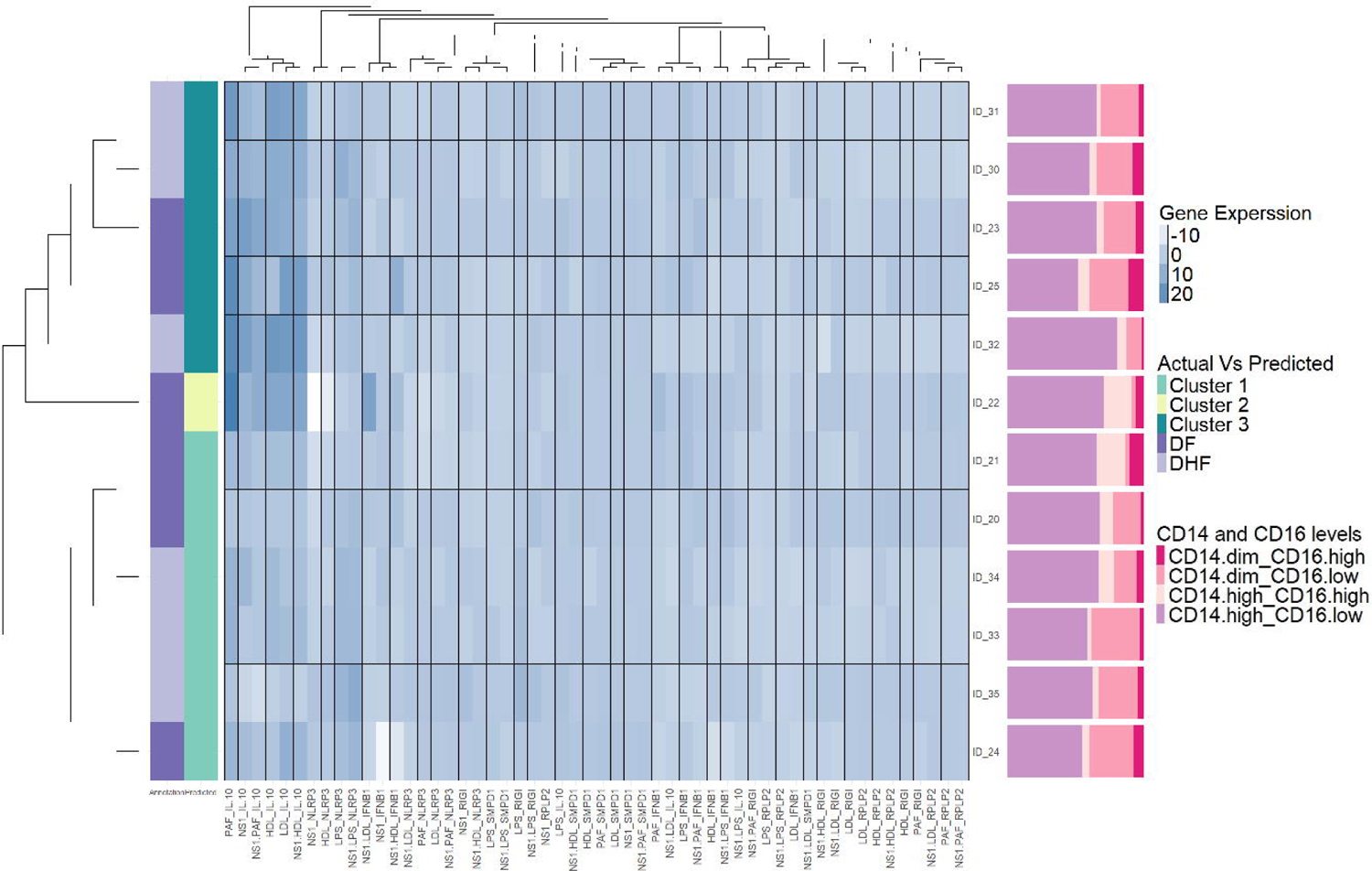
Patterns of expression of different genes in monocyte derived macrophages (MDMs) when co-cultured with NS1 and different lipid mediators. A heat-map was generated for expression of *RIGI, IL-10, NLRP3, IFN-*β*, SMPD1, RPLP2*, when MDMs were co-cultured from individuals with a history of DF (n=6) and DHF (n=6) with NS1 alone, different lipid mediators alone or with NS1 and different lipid mediators. Hierarchical cluster analysis with Euclidean distance calculations were used to differentiate clusters under different culture conditions of NS1 with different lipid mediators. The color intensity corresponds to the Z-score that indicates the number of standard deviations by which the normalized raw value was below or above. The differences of monocyte subpopulations identified of each individual is indicated in the Y axis.

As different monocyte subsets have shown to produce different cytokines in response to the DENV and other stimuli, we sought to investigate if those who had higher levels of *IL-10* expression compared to those with lower expression levels had a different composition of monocytes. To identify different monocyte subsets, PBMCs in those gene expression analysis was carried out were gated as follows: CD14^high^ CD16^low^, CD14^high^ CD16^high^, CD14^dim^ CD16^low^ and CD14^dim^ CD16^high^. The predominant monocyte subpopulation was the classical monocytes (CD14^+^CD16^−^), followed by non-classical monocytes (CD14^dim^CD16^+^), and intermediate monocytes (CD14^+^CD16^+^). We did not observe any significant differences in the proportion of different monocyte sub-populations in those who had high expression of *IL-10* (cluster 1) compared to those with lower *IL-10* expression (cluster 2).

### Differential gene expression in those with past DF and DHF when NS1 was co-cultured with different lipid mediators

We then proceeded to differential gene expression in those with past DF and DHF and constructed a volcano plot to identify significant differences between these two groups (Figure 3A). We identified significant differences in expression of *RPLP2*, when monocyte-derived macrophages of those with past DF or DHF were co-cultured with PAF alone, PAF+NS1 or NS1+LDL.

**Figure 3.**
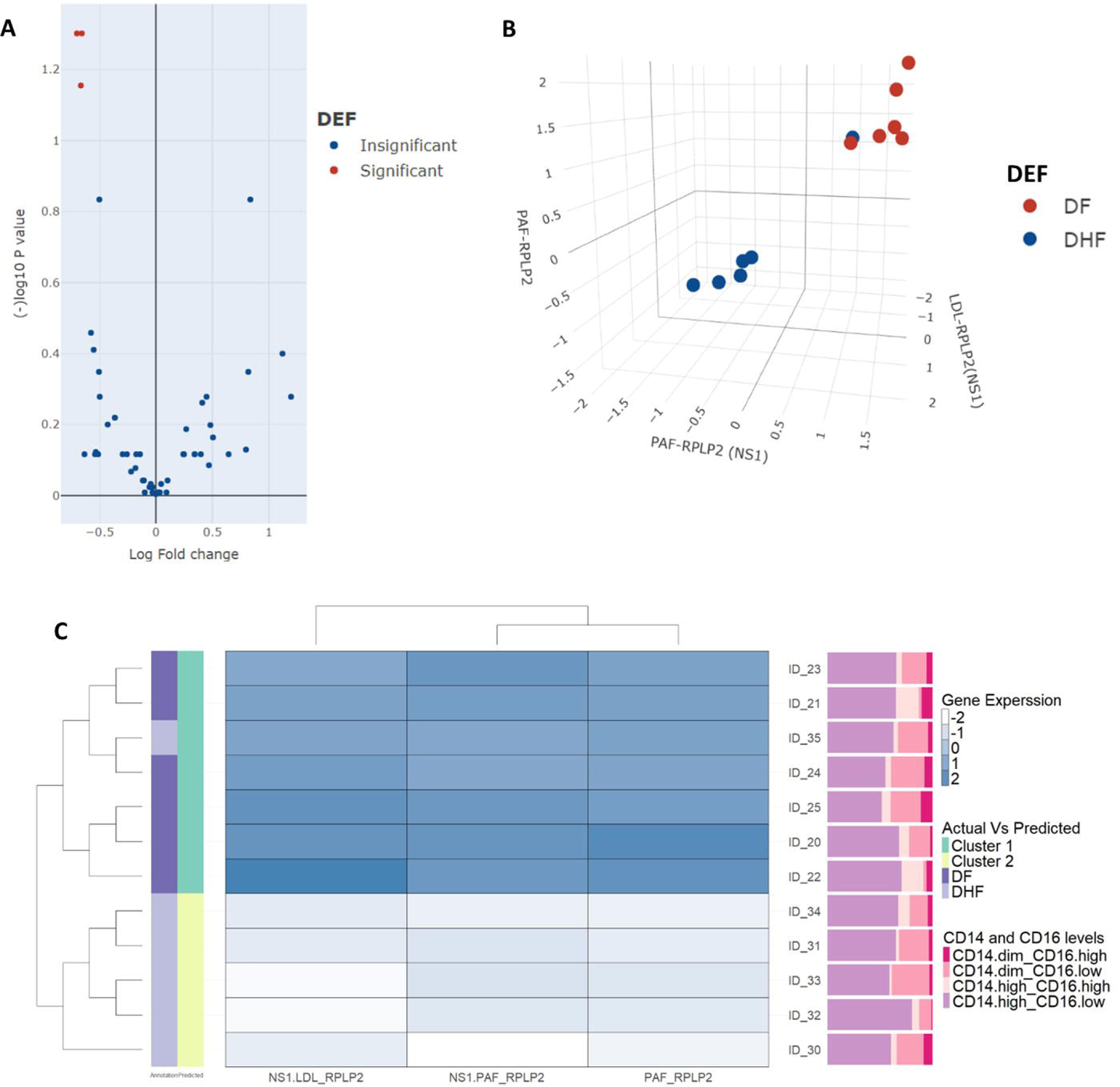
Differences in gene expression patterns in monocyte derived macrophages (MDMs) when co-cultured with NS1 and different lipid mediators in those with past DF and DHF. A volcano plot was generated to identify significant differences in gene expression in MDMs in those with past DF (n=6) and past DHF (n=6), with NS1 alone, different lipid mediators alone or with NS1 and different lipid mediators (A). Significant differences in *RPLP2* gene expression were observed when MDM2s of those with past DHF were co-cultured with PAF, NS1 and PAF and NS1 and LDL, which are also displayed in a 3D plot view with individuals with DHF (blue) and DF (red) (B). A heatmap was generated for differential expression of *RPLP2* identified by the volcano plot in those with past DF and DHF. The differences in monocyte subpopulations identified of each individual is indicated in the Y axis (C).

The differences in expression levels of *RPLP2* under these three culture conditions were then used to generate a 3D scatter plot. Furthermore, with the three features, the classifier model was created using 50% of the dataset and was found to predict the differences in the two biological groups (past DF and DHF) with 100% accuracy using our dataset. To avoid the bias by using our internal dataset, we then used the remaining 50% of the dataset as the external validation dataset, which gave the accuracy of the model as 68%. The generated 3D scatter plot and heatmaps using this model clearly distinguished those with past DF and DHF, except for one individual with past DHF who was also classified under the past DF group (Figure 3B and 3C).

## Discussion

In this study we show that primary monocyte-derived macrophages produced significantly higher levels of IL-β when NS1 was co-cultured with LDL, but not HDL. In fact, IL-β levels in culture supernatants were lower when the macrophages were co-cultured with HDL and NS1, rather than HDL alone. Thuse observations are contrary to those made by Benfrid et al, where they saw significantly higher levels of proinflammatory cytokines when macrophages were co-cultured with NS1 and HDL [5]. In our experiments, we used the exact conditions, and concentrations of HDL except the NS1 concentrations (NS1 concertation being 10 µg/ml vs our concentrations of 2µg/ml). For instance, Libraty *et al* reported NS1 antigen levels of ≥600ng/ml (0.6µg/ml) were associated with progression to DHF [23], Allonso *et al* reported a median NS1 antigen concentrations varying from 22.6 ng/mL to 36.8 ng/mL based on day of illness [1], and Nunes *et al* reported mean levels of 4.72 ng/mL and 4.92 ng/mL in those with primary and secondary dengue [28]. Although the NS1 concentrations used here was also higher than levels found in patients with acute dengue, we wished to use values that are closer to those reported in those with severe dengue [23]. Therefore, the differences between NS1 used in the experiments could account for the differences in results.

Previously we showed that primary human monocyte responses to all four serotypes of the DENV varied based on past dengue infection, with those with past DHF producing higher viraemia and proinflammatory cytokines in culture supernatants [21]. In this study, we observed that macrophages in individuals with past DHF produced significantly higher levels of IL-β, when they were co-cultured with NS1 and PAF. Furthermore, using volcano plots when we evaluated differential gene expression of macrophages in those with past DF and DHF we found that macrophages of individuals with past DHF, were significantly more likely to down-regulate *RPLP2* expression when co-cultured with PAF, NS1 and PAF and NS1 and LDL. The *RPLP2* gene encodes for 60S subunit of the ribosomes and is responsible for the elongation step of protein biosynthesis. RPLP2 has many functions including regulating glycolysis, cell proliferation, regulation of the MAPK1/ERK2 signaling pathway, regulation of generation reactive oxygen species (ROS), endoplasmic reticulum stress and also binds to TLR4 [2,17,42]. Furthermore, RPLP2 was shown to be required for many flaviviruses such as dengue, yellow fever and zika virus [9]. DENV replication results in accumulation of ROS and proinflammatory responses [14]. The unfolded protein response, which is induced as a result of endoplasmic reticulum stress in altered in DENV infection, resulting in altered autophagy and cellular immune responses [29]. High levels of expression of RPLP2 reduced accumulation of ROS and reduced ER stress and regulates the unfolded protein response [2,17]. Therefore, it is interesting to note that *RPLP2* gene expression was significantly down-regulated in macrophages of individuals with past DHF, when co-cultured with PAF, NS1 and PAF or NS1 and LDL. These differences observed in response to NS1 and different lipids in those with varying severity of past dengue, could be due to certain genetic or epigenetic changes in monocytes/macrophages of such individuals, which predispose them to have an altered response to NS1 and the DENV.

Although elevated IL-10 is associated with severe disease in many viral infections including dengue [24,31], compared to illnesses such as severe COVID-19, the IL-10 levels in those who have DHF are several folds higher [12]. PAF, NS1 and PAF, HDL and NS1 and LDL and NS1 combinations led to a significant upregulation of *IL-10* gene expression in one cluster of individuals, while the other cluster showed no upregulation or down regulation. These two clusters, which differed on *IL-10* expression levels did not associate with past dengue disease severity. As different monocyte subsets produce different cytokine profiles to inflammatory stimuli, we investigated the different monocyte subpopulations in those who had high *IL-10* expression vs normal expression levels and compared monocyte subpopulations in those with past DF and DHF. We did not find any difference in the monocyte subpopulation proportions in those who had high *IL-10* expression compared to no change in *IL-10* expression when co-cultured with NS1 and other lipids. Therefore, rather than differences in monocyte subpopulations, other mechanisms that lead to increased *IL-10* gene transcription could be responsible for these differences observed between the two clusters.

In summary, to delineate if NS1 and lipid interactions alters the pathogenicity of NS1, we investigated their effects on primary human monocyte derived macrophages. We found that NS1 and LDL led to increased production of IL-β, while significantly lower expression of *RPLP2* gene was observed in the macrophages of those with past DHF, when co-cultured with PAF, PAF and NS1 and NS1 and LDL. As RPLP2 is important in DENV replication and in regulating cellular stress responses and immune responses, it would be important to further explore how differential expression of RPLP2 could lead to disease pathogenesis.

## Data Availability

All data is available within the manuscript and supporting files.

## Acknowledgements

We are grateful to the NIH, USA (grant number 5U01AI151788-02), Accelerating Higher Education Expansion and Development (AHEAD) Operation of the Ministry of Higher Education funded by the World Bank, and the UK Medical Research Council.

---

**Supplementary Figure 1.** Monocyte phenotype example results of an individual. A.) Singlets were selected from the population of gated monocytes. B.) Monocyte subsets were selected from the live cell population and CD14^high^ CD16^low^, CD14^high^ CD16^high^, CD14^dim^ CD16^low^ and CD14^dim^ CD16^high^ phenotype percentages were obtained.

## Notes

### Competing Interest Statement

The authors have declared no competing interest.

