## Supplementary figures and images for "Dengue NS1 interaction with lipids alters its pathogenic effects on monocyte derived macrophages"

### Supplementary figure

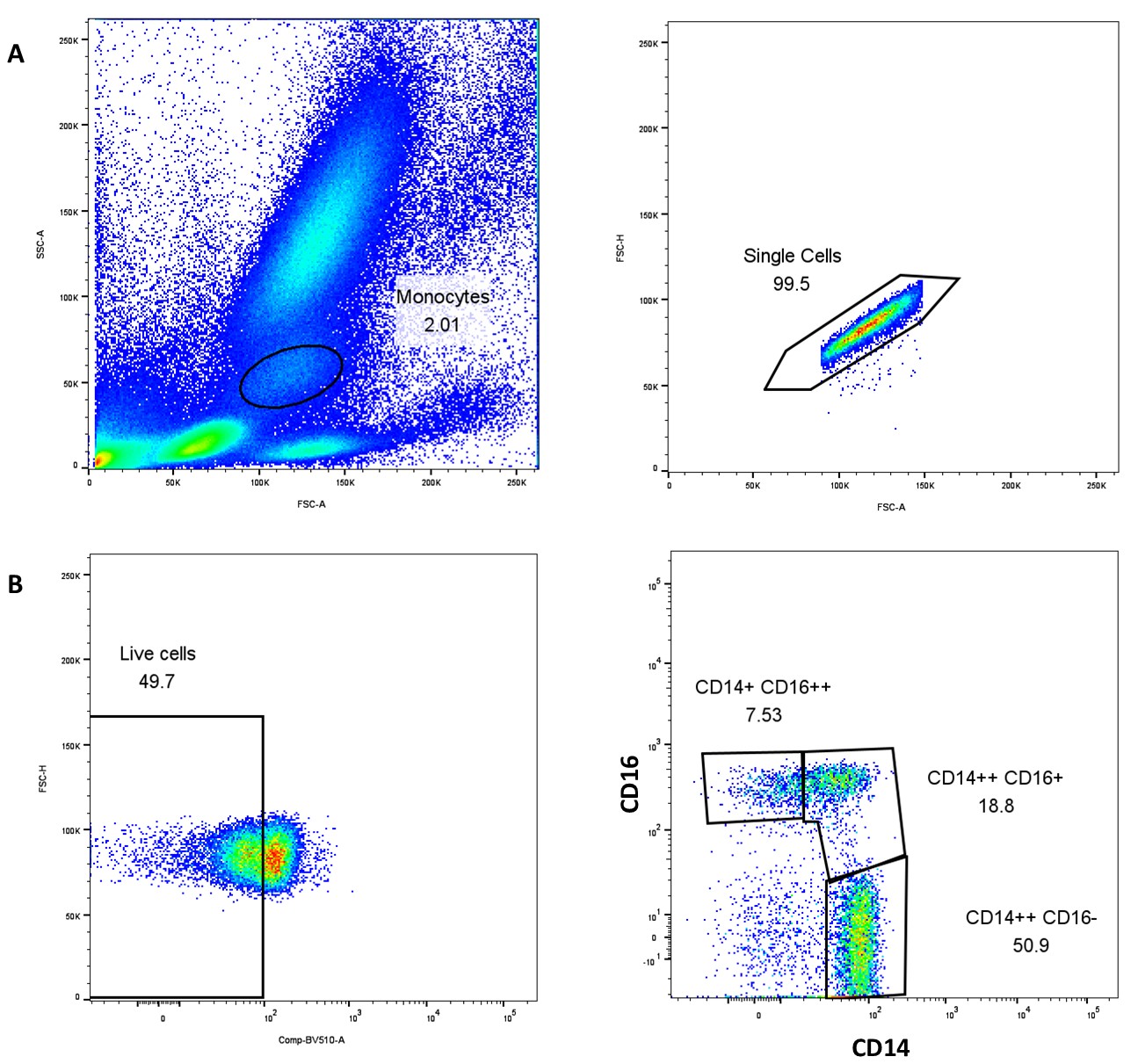
