## Supplementary data for "Dengue NS1 interaction with lipids alters its pathogenic effects on monocyte derived macrophages"

**Supplementary Table 1:** Past dengue disease severity details and the year of experiencing dengue

| Category | Sample ID | Age | Year of infection |
| --- | --- | --- | --- |
| DF | 20 | 25-30 | asymptomatic |
|  | 21 | 25-30 | 2017 (OPD) |
|  | 22 | 25-30 | 2017, 2019 |
|  | 23 | 40-45 | 2017 |
|  | 24 | 30-35 | 2019 |
|  | 25 | 50-55 | asymptomatic |
|  | 26 | 30-35 | asymptomatic |
|  | 27 | 25-30 | asymptomatic |
| DHF | 30 | 25-30 | 2005 |
|  | 31 | 25-30 | 2013, 2021 |
|  | 32 | 30-35 | 2017 |
|  | 33 | 25-30 | 2000 |
|  | 34 | 35-40 | 2022 |
|  | 35 | 30-35 | 2019 |
|  | 36 | 20-25 | 2023 |
|  | 37 | 25-30 | 2007 |
